# Co-Design of the VIRtual physiotherapist-led Triage of referrals to shoUlder surgEons (VIRTUE) model of care: a study protocol

**DOI:** 10.64898/2026.11.05.26367222

**Authors:** Andrew R Gamble, Giovanni E Ferreira, Chris G Maher, Samuel Whittle, Christos Avdalis, Ian A Harris, Aili Langford, Christopher M Williams, Karen Tambree, Leanne Hassett, Abby Haynes, Peter Malliaras, David B Anderson, Joshua R Zadro

## Abstract

**Background:** Rotator cuff-related shoulder pain accounts for up to 85% of shoulder pain. Guidelines recommend 6 months of non-surgical care before considering surgery, yet many are referred to a surgeon before trying it. High referral rates to public hospital shoulder surgery clinics in Australia mean many patients wait 1-2 years to see a surgeon, during which symptoms may become more complex and costly to manage. A promising solution is the VIRtual physiotherapist-led Triage of referrals to shoUlder surgEons (VIRTUE) model of care, in which physiotherapists screen new referrals and determine suitability for non-surgical care or surgical review. However, no such model has been co-designed to incorporate virtual care nor evaluated in a randomised controlled trial.

**Aims:** The broad aim of this study is to co-design the VIRTUE model of care ready for evaluation in a randomised controlled trial conducted in public hospital shoulder surgery clinics in Australia. Specifically, this study has two aims (two stages): i) explore stakeholders’ perceptions of the feasibility, acceptability and implementation of the VIRTUE model of care for adults with rotator cuff-related shoulder pain who are referred to public hospital shoulder surgery clinics; and ii) co-design the VIRTUE model of care so that it is user-centred and acceptable to patients, clinicians and key stakeholders.

**Methods:** Stage 1 is a qualitative descriptive study using semi-structured online interviews with patients, clinicians and key stakeholders, recruited using purposive, maximum variation sampling. Interviews will explore experiences of current care, perceptions of the proposed model, and barriers and facilitators to implementation, guided by the Theoretical Framework for Acceptability and Consolidated Framework for Implementation Research. Stage 2 will use Experience-Based Co-Design methodology and involves three online workshops: separate workshops for patients, and for clinicians and key stakeholders, then a combined workshop. Workshops will explore Stage 1 findings, perceptions of the proposed model, and refinements needed to finalise it, with data mapped to the Framework for Reporting Adaptations and Modifications-Expanded.

**Conclusion:** Co-designing VIRTUE with patients, clinicians and key stakeholders will ensure it is user-centred, acceptable and feasible within Australian public health services, ready for evaluation in a randomised controlled trial. By facilitating timely and equitable access to evidence-based care, VIRTUE could improve outcomes for patients, ensure specialist services are available for those most in need, and improve health system performance.

## 1. Introduction

Shoulder pain is the third most common musculoskeletal complaint seen by primary care providers in Australia (e.g., general practitioners).^1^ It has a lifetime prevalence of up to 67% in the general population,^2^ and is estimated to affect 500 million people globally at any given time.^3^ Rotator cuff-related shoulder pain is the most common shoulder condition, accounting for up to 85% of cases of shoulder pain, and is the most burdensome shoulder condition in terms of affecting work and self-care activities.^4^ Guidelines for rotator cuff-related shoulder pain recommend 6 months of non-surgical care (e.g., activity advice, exercise therapy) before considering surgery,^5^ based on high-certainty evidence that surgery is not superior to placebo or non-surgical care. Despite this, ~20,000 surgeries are performed each year in Australia,^6^ costing ~$180 million.^7^

High referral rates to public hospital shoulder surgery clinics mean many clinics in Australia struggle to assess new non-urgent cases (i.e., not due to serious pathology) within 1-2 years.^8^ In many cases these patients have not tried non-surgical care, and while they wait to see a surgeon, they develop disabling symptoms which are more complex and costly to manage.^9^ This problem is exacerbated in rural and remote areas where orthopaedic waiting times are even longer than metropolitan areas.^10 11^

A potential solution to help patients receive guideline-recommended care sooner is the VIRtual physiotherapist-led Triage of referrals to shoUlder surgEons (VIRTUE) model of care. VIRTUE involves physiotherapists screening new referrals to shoulder surgery clinics and determining their suitability for a course of non-surgical care or surgical review. Similar models of care have been explored in some shoulder surgery clinics in Australia. For example, a retrospective study found an in-person physiotherapist-led triage service improved outcomes and satisfaction and reduced the wait list of a Sydney shoulder surgery clinic by 34%.^8^ However, no randomised controlled trial has evaluated the clinical and cost-effectiveness of this service in Australia or globally, and no one has explored a model developed using a formal co-design process. Co-design could promote shared ownership of the model and identify important service design characteristics vital to implementation and scale-up success (e.g., use of a virtual service to improve access for people in rural and remote areas, developing a patient-centred model, understanding site-specific workflow, contextual barriers, roles and responsibilities). Before rigorous evaluation of this model in a clinical trial, a user-centred and stakeholder-endorsed version must be co-designed to ensure feasibility and acceptability across rural, remote and metropolitan sites, and successful translation and scale-up if effective.

The co-design and evaluation of the VIRTUE model of care will be conducted across three phases. The first phase (Stage 1 of this study) will involve qualitative interviews to explore patients’, clinicians’ and key stakeholders’ perceptions of the proposed model of care. The second phase (Stage 2 of this study) will involve three online workshops with patients, clinicians and key stakeholders to finalise the model of care. Co-design is defined as ‘a collaborative approach that actively involves service users, providers, and other stakeholders in service design, ensuring those most affected have a meaningful role in shaping decisions’.^12^

^13^ The third phase will be the evaluation of the VIRTUE model of care in a randomised controlled trial to be conducted in Australian public hospital shoulder surgery clinics.

## 2. Aims

The broad aim of this study is to co-design the VIRTUE model of care ready for evaluation in a randomised controlled trial conducted in public hospital shoulder surgery clinics in Australia. Specifically, this study has two aims (two stages):

1. Explore stakeholders’ perceptions of the feasibility, acceptability and implementation of the VIRTUE model of care for adults with rotator cuff-related shoulder pain who are referred to public hospital shoulder surgery clinics; and
2. Co-design the VIRTUE model of care so that it is user-centred and acceptable to patients, clinicians and key stakeholders (e.g., department heads).

## 3. Methods

### 3.1 Stage 1: Qualitative study

This qualitative study will explore stakeholders’ perceptions of the feasibility, acceptability and implementation of the VIRTUE model of care for adults with rotator cuff-related shoulder pain who are referred to public hospital shoulder surgery clinics. It has been approved by The University of Sydney Human Research Ethics Committee (HREC) (2025/HE001266).

#### 3.1.1. Study Design

This is a qualitative descriptive study with in-depth, semi-structured interviews. Qualitative description aims to provide a synthesised description of stakeholder views and experiences on a healthcare topic which can inform practice.^14^

#### 3.1.2 Participants

We will recruit patients, clinicians and key stakeholders. Eligible participants need to be able to speak English in order to take part in the interviews. Eligible ***patients*** are adults living in Australia with lived experience of rotator cuff-related shoulder pain. Eligible ***clinicians*** are clinicians who are registered to practise in Australia (evidenced by current AHPRA registration) and who are involved in the management of people with rotator cuff-related shoulder pain and treated at least 5 patients with rotator cuff-related shoulder pain in the last 12 months. Examples include, but are not limited to: physiotherapists, shoulder surgeons, general practitioners, rheumatologists, psychologists, and exercise physiologists. ***Key stakeholders*** are those who work for an organisation that has an interest in healthcare for people with rotator cuff-related shoulder pain. Examples include, but are not limited to: heads of physiotherapy or orthopaedic departments at hospital sites in NSW, and representatives from the Australian Physiotherapy Association, Sydney Health Partners, and government funded insurance authorities.

#### 3.1.3. Recruitment

##### Patients

We will recruit patients via a brief advertisement on social media platforms X, LinkedIn and Facebook. The advertisement will briefly describe the study and provide a link to a welcome page of the survey providing more detail, as well as the Participation Information Statement and Consent Form. We will also use passive snowball recruitment to recruit patients, where participants will be asked to discuss the research with others who may be interested (and eligible) to participate. The potential participants will then be invited to contact the research team to discuss their involvement if they are interested.

##### Clinicians

We will recruit clinicians through social media or our existing network of clinician collaborators (e.g., contacts from the research team’s current or previous workplace) with a formal invitation letter, sent by post, fax or email, seeking an expression of interest from individual clinicians. We will also contact relevant professional organisations and seek their permission to advertise the study through their mailing lists or on their websites (e.g. Australian Physiotherapy Association, Australian Orthopaedic Association, Royal Australian College of General Practitioners, Rural Doctors Association of Australia, Australian Rheumatology Association), and use a passive snowballing recruitment technique by asking participating clinicians to refer or forward the invitation letter to their colleagues (or patients with rotator cuff-related shoulder pain). The potential participants will then be invited to contact the researchers to discuss their involvement if they are interested.

##### Key stakeholders

We will recruit key stakeholders through our existing network of clinician collaborators (e.g., contacts from the research team’s current or previous workplace) with a formal invitation letter, sent by post, fax or email, seeking an expression of interest to participate in the study. These key stakeholders may include, but are not limited to: heads of physiotherapy or orthopaedic departments, government bodies, insurance companies, professional associations, healthcare organisations, and consumer advocacy organisations.

We will also use passive snowball recruitment to recruit other key stakeholders, where participants will be asked to discuss the research with colleagues who may be interested (and eligible) to participate. The potential participants will then be invited to contact the researchers to discuss their involvement if they are interested.

##### Eligibility screening and consent

For all participant groups, potential participants will first read the Participant Information Statement and Consent Form and provide consent by clicking ‘Yes’ at the beginning of the online Pre-Interview Questionnaire. They will then complete screening questions to confirm eligibility, and those deemed eligible will answer the remaining questions in the same questionnaire (see Section 3.1.5).

#### 3.1.4. Sampling Method

We will use purposive sampling, a research strategy which supports the non-randomised, deliberate seeking and selection of specific types of participants.^15^ We will aim for maximum variation so that our sample includes individuals who vary from each other as much as possible.^16^ This will allow us to triangulate data on the topic from diverse perspectives. Participant characteristics for maximum variation sampling include age, gender, location (metropolitan/regional/rural), country of birth, length and severity of patients’ symptoms, their health literacy, and clinician confidence in using telehealth.^17^ Given that we may seek to fill gaps in our data as recruitment progresses (e.g., by targeting people living in regional/rural areas if we have mostly heard from metropolitan-participants), the Participant Information Sheet will make clear that even if eligible, participation is not guaranteed.

We anticipate interviewing approximately 12-16 patients and a combination of 12-16 clinicians and key stakeholders. Participants will be selected until data saturation is reached, i.e., the point at which the researchers judge that no new perspectives relevant to the research questions are being identified in interviews.^18,19^ However, if recruitment is slow we may cease data collection before this if we judge we have sufficient rich information about diverse perspectives to guide design considerations regarding the feasibility, acceptability and implementation of the proposed model of care.^20^

#### 3.1.5. Data Collection

##### Pre-Interview Questionnaire

The Pre-Interview Questionnaire is the online survey which includes the screening questionnaire and will facilitate purposive sampling. (Appendix 1) It poses questions based on personal and professional factors to allow diversity in participant selection and transparency of non-identifiable characteristics of study participants. This gathers information on eligibility and demographic data, which will be used to satisfy sampling methods, and to inform the processes of data analysis. This is also where the participant can choose to submit their contact details.

##### Semi-Structured Interviews

Selected eligible participants will be contacted to participate in a semi-structured in-depth interview to explore their perceptions on the feasibility, acceptability and implementation of the VIRTUE model of care in public hospital shoulder surgery clinics. In summary, the VIRTUE model of care (Figure 1) involves physiotherapists screening new referrals to shoulder surgery clinics and determining their suitability for a course of non-surgical care or surgical review. Semi-structured interviews will employ open-ended questions which focus on follow-up queries of ‘why’ and ‘how’.^21^ This suits the objective of this project well, due to the heavy focus of the research aim on exploration. Questions in the interview guide have been informed by constructs in two relevant implementation and evaluation frameworks: the Theoretical Framework for Acceptability (TFA)^22^ and the Consolidated Framework for Implementation Research (CFIR).^23^ As is usual in qualitative research, the questions may be modified as interviews progress to take account of new learning (e.g. rephrasing a question so it is more meaningful or asking an additional question about an emergent topic raised by previous interviewees).^24^

**Figure 1.**
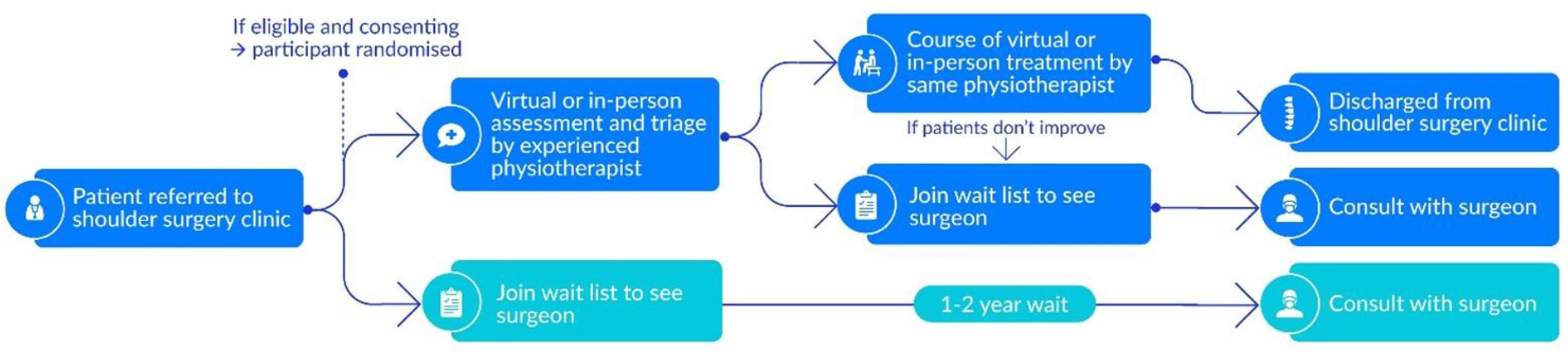
Virtual physiotherapist-led Triage of referrals to shoulder surgeons (VIRTUE) model of care

Eligible participants will be contacted to organise a suitable time, and the interview will take place over Zoom, lasting between 30-60 minutes. The researcher conducting the interviews will maintain a running memo in which they make reflective notes on key learnings following each interview. This reflection will include critical review of the interview guide (Appendix 2) and any changes made as interviews progress.

#### 3.1.6. Data Analysis

Questionnaire data will be summarised as means (standard deviations) or frequencies and proportions when appropriate. Qualitative data analysis and collection will be completed concurrently. Interviews will be transcribed using Zoom’s transcription feature, which will then be checked over by a member of the research team for accuracy. The researchers conducting the participant interviews will be responsible for the analysis of the data which will be reviewed by a senior researcher before being reviewed by the wider research team.

Transcripts will be imported into the qualitative software program NVivo and analysed using a qualitative descriptive approach which aims to present a straightforward, low inference account of participants’ views that stays close to the data and is written up so that the results can be of applied use.^25^ The interview guide (Appendix 2) will aid in mapping emerging themes to the Theoretical Framework for Acceptability (TFA)^22^ and Consolidated Framework for Implementation Research (CFIR).^23^ The TFA consists of seven constructs (affective attitude, burden, perceived effectiveness, ethicality, intervention coherence, opportunity cost and self-efficacy) which can inform the evaluation of the acceptability of the VIRTUE model of care.^22^ The CFIR will provide a framework for the assessment of contextual factors such as barriers and facilitators to the potential feasibility of the new model of care.^23^

Early in the data collection process, two researchers will develop an initial coding frame based on the a priori concepts targeted by the interview questions in relation to participants’ views about acceptability and feasibility of the proposed model of care and suggestions for improvements. Key issues identified in the running memo will also inform this process. Inductive codes will be added to capture emergent concepts in the data which are relevant to deepening our understanding of the research topic and may inform the subsequent co-design process. The researchers will trial this coding frame on two transcripts for each stakeholder group, working collaboratively to refine the coding frame and develop concordance about the scope and content of each code as it evolves. One researcher will code further transcripts with continued revisions to the coding frame to capture variations in the data, if required.

When approximately half of the transcripts have been coded and an initial overview of key learnings has been developed, a workshop will be held with other research team members with multidisciplinary backgrounds who will review the analysis that has been performed, with the goal of refining findings and identifying key learning that can most usefully inform the next phase of work. This will inform subsequent data analysis.

Strategies to enhance research rigour will be used throughout. These include purposive sampling for maximum variation; constant comparative analysis that moves iteratively between data collection, analysis and critical review of findings; reflective memoing; and triangulation of different perspectives from stakeholders (data triangulation) and the multidisciplinary investigator team (researcher triangulation).^26^

### 3.2. Stage 2: Co-design workshops

This co-design study will develop the VIRTUE model of care so that it is user-centred and acceptable to patients, clinicians and key stakeholders (e.g., patients with rotator cuff-related shoulder pain, shoulder surgeons, physiotherapists, department heads). It has been approved by The University of Sydney Human Research Ethics Committee (2025/HE001334).

#### 3.2.1. Study Design

Stage 2 will use Experience-Based Co-Design (EBCD) methodology. EBCD is a participatory approach primarily used for improving healthcare quality.^27^ This multi-stage process engages patients and healthcare staff in identifying ways to enhance service experiences and improve care delivery.^27^ Findings from the Stage 1 qualitative study (Ethics reference: 2025/HE001266) will inform the Stage 2 co-design of the model of care.

#### 3.2.2. Participants

Eligibility criteria for patients and clinicians are the same as in Stage 1 (see Section 3.1.2).

#### 3.2.3. Recruitment

Recruitment for Stage 2 will be the same as for Stage 1 (see Section 3.1.3). However, select patients, clinicians and key stakeholders who took part in the Stage 1 qualitative study will also be invited to participate in the co-design workshops.

#### 3.2.4. Sampling Method

Sampling methods used will be purposive sampling, namely criterion and maximum variation sampling. Maximum variation sampling will enable a sample in which individuals vary from each other as much as possible^16^, enabling a broader view of the topic. Despite not including theoretical sampling, the adherence to purposive sampling will still allow the non-randomised, deliberate seeking and selection of participants.^15^ The Participant Information Sheet will make clear that even if eligible, participation is not guaranteed.

Anywhere between 6 to 12 participants from each representative group (i.e., patient group, and clinician and key stakeholder group) will be selected for the co-design workshops. This will allow for active participation, diverse perspectives, and manageable group discussions.^28^ The exact number of participants cannot be determined as it will vary depending on the workshops’ complexity and the desired depth of discussion required,^29 30^ but the proposed sample size range is commonly used and reported in co-design studies with similar aims (i.e., co-design to develop/adapt models of care).

#### 3.2.5. Consent

Sufficient information will be provided to participants regarding the study via a Participant Information Statement. Formal written consent to complete the survey and willingness to participate in the study will be provided using tick boxes in the pre-screening questionnaire on Qualtrics survey software. Ticking the appropriate box will acknowledge the participants have reviewed the appropriate Participant Information Statement and Participant Consent Form. That is, participants who are considered to be eligible after filling in the pre-workshop questionnaire (Appendix 1) and who submit the online questionnaire providing additional information will have consented to entering the study. Participant consent will also be confirmed verbally by the workshop facilitator.

#### 3.2.6. Data Collection

Selected eligible participants will be invited to attend three structured, group-based workshops to co-design the VIRTUE model of care. These will comprise separate workshops for patients (Workshop 1) and for clinicians and key stakeholders (Workshop 2), followed by a combined workshop (Workshop 3). Workshops will be held online via Zoom, with an anticipated duration of 1-2 hours each. In Workshops 1 and 2, the Stage 1 qualitative study results will be reviewed and discussed before participants are asked to reflect and comment on the proposed VIRTUE model of care. The findings from each of these workshops will be analysed before developing a revised model of care that will be presented and discussed in Workshop 3. Following Workshop 3, further refinements will be made before the model of care is finalised.

The workshops will be facilitated online via Zoom by a researcher experienced in co-design. The facilitator will guide the workshops based on the responses and a series of activities to prompt responses. (Appendix 2)

##### Workshop 1 Objective

Gather patient perspectives on the qualitative study findings and proposed model of care.

##### Workshop 2 Objective

Gather clinicians’ and key stakeholders’ perspectives on the qualitative study findings and proposed model of care.

##### Workshop 3 Objective

Bring together patient, clinician and key stakeholder feedback to finalise the model of care.

Data collection will be completed during each workshop through Zoom audio and video recording and research team members simultaneously taking notes on key points from each workshop. Workshops will be transcribed using the automated transcription feature in Zoom, which will then be checked over by a member of the research team to ensure accuracy before deleting the audio and video recordings.

#### 3.2.7. Data Analysis

Analysis will occur immediately after each end-user workshop; data will be analysed by mapping experiences against an adaptation framework - The Framework for Reporting Adaptations and Modifications-Expanded (FRAME).^31^ The FRAME provides a structure by categorising the information collected during the workshops and document adaptations to the VIRTUE model of care that are necessary. Summary reports will be produced for each workshop and a final outline of the VIRTUE model of care that will be confirmed through consensus by participants in the third co-design workshop.

## Data Availability

All data produced in the present study are available upon reasonable request to the authors.

