## Appendix 1 for "Co-Design of the VIRtual physiotherapist-led Triage of referrals to shoUlder surgEons (VIRTUE) model of care: a study protocol"

**Appendix 1. Pre-interview questionnaires and pre-workshop questionnaires**

### Co-Design of the VIRtual physiotherapist-led Triage of referrals to shoUlder surgEons (VIRTUE) model of care: a study protocol

**Stage 1: Qualitative study pre-interview questionnaire first page (landing page):**

Thank you for your interest in our study. Our team of researchers from The University of Sydney are conducting a research study to understand people's perceptions about a new model of care that aims to reduce waiting times to see a shoulder surgeon for rotator cuff-related shoulder pain in Australian public hospitals. We are interested in understanding your views on the new VIRtual physiotherapist-led Triage of referrals to shoUlder surgEons (VIRTUE) model of care as we look to develop it further.

If you agree to participate in this study, you will be asked to complete a short survey about you (10 minutes). You may then be asked to attend a one-on-one interview, but this is not guaranteed. We aim to interview people with diverse characteristics and opinions, so whether you are selected for an interview will depend on whom we have already interviewed. Interviews will be conducted remotely via Zoom and will between 30-60 minutes.

First, there are a few screening questions to confirm your eligibility...

**Stage 2: Co-design workshops pre-workshop questionnaire first page (landing page):**

Thank you for your interest in our study. Our team of researchers from The University of Sydney are conducting a research study to understand people's perceptions about a new model of care that aims to reduce waiting times to see a shoulder surgeon for rotator cuff-related shoulder pain in Australian public hospitals. We are interested in understanding your views on the new Co-Design of the VIRtual physiotherapist-led Triage of referrals to shoUlder surgEons (VIRTUE) model of care as we look to refine it further before evaluating it in a randomised controlled trial.

If you agree to participate in this study, you will be asked to complete a short survey about you (10 minutes). You may then be asked to attend two workshops, but this is not guaranteed. We aim to involve people with diverse characteristics and opinions, so whether you are selected will depend on whom we have already chosen. Workshops will be conducted remotely via Zoom and will between take between 1-2 hours each.

First, there are a few screening questions to confirm your eligibility...

**Appendix 1. Pre-interview questionnaires and pre-workshop questionnaires**

**Stage 1 and Stage 2 questionnaires are identical and presented below:**

**[Pre-screening questionnaire - Patients]**

1. Do you live in Australia?
   - ☐ Yes
   - ☐ No [screen out]

2. Do you speak English?
   - ☐ Yes
   - ☐ No [screen out]

3. Are you a person with rotator cuff-related shoulder pain?
   - ☐ Yes
   - ☐ No [screen out]

**[Pre-screening questionnaire – Clinicians]**

1. Do you live in Australia?
   a. Yes
   b. No [screen out]

2. Are you a clinician who manages people with rotator cuff-related shoulder pain? *(i.e. you have reviewed at least 5 patients with* rotator cuff-related shoulder pain *in the last 12 months)*
   a. Yes
   b. No [screen out]

**[Pre-screening questionnaire – Key Stakeholders]**

1. Do you live in Australia?
   a. Yes
   b. No [screen out]

2. Are you a key stakeholder?
   (e.g. someone working for a government agency, health insurance company, healthcare organisation, consumer advocacy organisation or others)?

**Appendix 1. Pre-interview questionnaires and pre-workshop questionnaires**

a. Yes
b. No [screen out]

It appears you are eligible for the study.

Please read the Participant Information Statement below to ensure that you understand what is involved in this study. By selecting 'yes' below, you are indicating that you have read and understood the [link to Participant Information Sheet] *(click to download)*, and that you consent to participate in this study.

[Participant Consent Form and post-consent questionnaire to be shown only to those who were not screened out]

[Insert Participant Consent Form]

In giving my consent, I confirm that that:

- ☐ Yes, I would be happy to complete the survey and participate in this study
- ☐ No, I would prefer not to complete the survey or participate in this study

Contact details:

*Please provide your best contact details so a researcher from the University of Sydney can contact you to arrange the interview time:*

Name: ____________________________

Email: ________________________________

Best contact telephone number: ________

**When are the best times to schedule the interview for you?**

| | **Monday** | **Tuesday** | **Wednesday** | **Thursday** | **Friday** |
|---|---|---|---|---|---|
| **6 – 8am** | | | | | |
| **8 – 10am** | | | | | |
| **10 – 12pm** | | | | | |
| **12 – 2pm** | | | | | |

**Appendix 1. Pre-interview questionnaires and pre-workshop questionnaires**

| |
|---|
| **2 – 4pm** |
| **4 – 6pm** |
| **6 – 8pm** |

**[The questions below will appear to eligible and consenting patient participants]**

We would now like to ask a few questions about yourself.

3. What is your age (in years): ____

4. What was your sex assigned at birth:
   - ☐ Male
   - ☐ Female
   - ☐ Another term (please specify)

5. What is your postcode: _____

6. What is your country of birth? (drop down menu with all countries provided)

7. What option best describes your highest level of education?
   a. Bachelor Degree or Higher
   b. Advanced Diploma or Diploma
   c. Certificate III or IV (including Trade Certificate)
   d. Year 12 or equivalent
   e. Year 11 or below
   f. Did not go to school

8. Do you have private health insurance?
   a. Yes
   b. No

9. What is the total income your household usually receives (pre-tax)?
   a. $6,000 or more per week ($312,000 or more per year)
   b. $3,000 - $5,999 per week ($156,000 - $311,999 per year)
   c. $1,600 - $2,999 per week ($83,200 - $155,999 per year)
   d. $800 - $1,599 per week ($41,600 - $83,199 per year)
   e. $1 - $799 per week ($1 - $41,599 per year)
   f. Nil or negative income

10. Have you ever seen a surgeon for your rotator cuff-related shoulder pain before?
   a. No

b. Yes

11. Have you ever had surgery for your rotator cuff-related shoulder pain before?
    a. No
    b. Yes

12. [Follow-up question that will only appear to people answering yes to the previous question] Which surgery did you have?
    a. Subacromial decompression surgery
    b. Rotator cuff repair surgery (with or without decompression)
    c. Other surgery (please specify)
    d. Unsure

[Numerical Pain Rating Scale]

13. On a scale of 0 to 10, where 0 represents no pain at all and 10 represents the worst possible pain, please select the number that best describes your rotator cuff-related shoulder pain over the last 4 weeks?

0 1 2 3 4 5 6 7 8 9 10
No pain at all Worst possible pain

[Shoulder Pain and Disability Index (SPADI) Outcome measure]

14. When your shoulder hurts, you may find it difficult to do some of the things you normally do.

Please place a mark on the line that best represents your experience during the last week attributable to your shoulder problem.

Pain scale:
How severe is your pain?
Circle the number that best describes your pain where: 0 = no pain and 10 = the worst pain imaginable.
1. At its worst? 0 1 2 3 4 5 6 7 8 9 10
2. When lying on the involved side? 0 1 2 3 4 5 6 7 8 9 10
3. Reaching for something on a high shelf? 0 1 2 3 4 5 6 7 8 9 10
4. Touching the back of your neck? 0 1 2 3 4 5 6 7 8 9 10
5. Pushing with the involved arm? 0 1 2 3 4 5 6 7 8 9 10

Disability scale:
How much difficulty do you have?
Circle the number that best describes your experience where: 0 = no difficulty and 10 = so difficult it requires help.

**Appendix 1. Pre-interview questionnaires and pre-workshop questionnaires**

1. Washing your hair? 0 1 2 3 4 5 6 7 8 9 10
2. Washing your back? 0 1 2 3 4 5 6 7 8 9 10
3. Putting on an undershirt or jumper? 1 2 3 4 5 6 7 8 9 10
4. Putting on a shirt that buttons down the front? 1 2 3 4 5 6 7 8 9 10
5. Putting on your pants? 1 2 3 4 5 6 7 8 9 10
6. Placing an object on a high shelf? 1 2 3 4 5 6 7 8 9 10
7. Carrying a heavy object of 10 pounds (4.5 kilograms) 0 1 2 3 4 5 6 7 8 9 10
8. Removing something from your back pocket? 1 2 3 4 5 6 7 8 9 10

Finally, would you like to receive a summary of the study results via email?

- ☐ Yes
- ☐ No

**We thank you for your time spent taking this survey. Your response has been recorded.**

**[The questions below will appear to eligible and consenting clinician participants]**

We would now like to ask a few questions about yourself.

1. What is your age (in years): ____

2. What is your sex assigned at birth:
    - ☐ Male
    - ☐ Female
    - ☐ Another term (please specify)

3. Which country did you attain your qualification as a health professional? (drop down menu with all countries provided)

4. What is your clinical background?
    - ☐ Medicine [those choosing this option will be asked a follow-up question to include specialty]
        - o General practice
        - o Rheumatology
        - o Neurosurgery
        - o Orthopaedic surgery
        - o Anaesthesiology
        - o Occupational and environmental medicine
        - o Emergency medicine
        - o Rehabilitation medicine
        - o Other (please specify)
    - ☐ Physiotherapy
    - ☐ Psychology

**Appendix 1. Pre-interview questionnaires and pre-workshop questionnaires**

- ☐ Exercise physiology
- ☐ Pharmacy
- ☐ Chiropractic
- ☐ Osteopathy
- ☐ Nursing
- ☐ Other (please specify)

5. What is the postcode of your most common place of practice? *(If you work across multiple, choose the one where you see the most patients with rotator cuff-related shoulder pain).*

6. Which State or Territory do you spend the most time practicing in?
   - ☐ New South Wales
   - ☐ Victoria
   - ☐ Queensland
   - ☐ South Australia
   - ☐ Western Australia
   - ☐ Tasmania
   - ☐ Northern Territory
   - ☐ Australian Capital Territory

7. Which clinical setting have you spent the most time practicing in?
   - ☐ Private practice
   - ☐ Public hospital
   - ☐ Private hospital
   - ☐ Other (please specify)

8. How long have you been practising for (in years)? ___

9. What is the postcode of your most common place of practice? ___

10. On average, how many patients with rotator cuff-related shoulder pain do you manage/review per week?
    - ☐ At least 5
    - ☐ 6-10
    - ☐ 11-20
    - ☐ 21-50
    - ☐ >50

Finally, would you like to receive a summary of the study results via email?

- ☐ Yes
- ☐ No

**We thank you for your time spent taking this survey. Your response has been recorded.**

**Appendix 1. Pre-interview questionnaires and pre-workshop questionnaires**

**[The questions below will appear to eligible and consenting key stakeholder participants]**

We would now like to ask a few questions about yourself.

3. What is your age (in years): ____

4. What is your sex assigned at birth:
    - ☐ Male
    - ☐ Female
    - ☐ Another term (please specify)

5. What is your country of birth? (drop down menu with all countries provided)

6. Which healthcare stakeholder do you primarily work for/represent?
    - ☐ Government agency
    - ☐ Health insurance company
    - ☐ Healthcare organisation
    - ☐ Consumer advocacy organisation
    - ☐ Other (please specify)

Finally, would you like to receive a summary of the study results via email?
- ☐ Yes
- ☐ No

**We thank you for your time spent taking this survey. Your response has been recorded.**
