## Appendix 2 for "Co-Design of the VIRtual physiotherapist-led Triage of referrals to shoUlder surgEons (VIRTUE) model of care: a study protocol"

**Appendix 2. Interview and workshop guides**

### Co-Design of the VIRtual physiotherapist-led Triage of referrals to shoUlder surgEons (VIRTUE) model of care: a study protocol

#### INTERVIEW GUIDE – mapped to the Theoretical Framework for Acceptability (TFA) and Consolidated Framework for Implementation Research (CFIR)

*Note: The topics below will serve as an outline to guide the interview.*

##### Stage 1: Qualitative study interview guides:

**Introduction**
*Introduce yourself to the informant.*

*Thank you for agreeing to participate in today's interview. We are aiming to take only 30-60 minutes of your time today. This interview is voluntary and at any point of time you are free to withdraw.*

*The purpose of this interview is to learn about your experience of rotator cuff-related shoulder pain in our health system and to explore your views on different ways of managing care for people with rotator cuff-related shoulder pain, particularly people with persistent rotator cuff-related shoulder pain who have been referred to see a shoulder surgeon.*

*[Present infographic on model of care while explaining the below]*

*In the current model of care, an increasing number of referrals has led to many shoulder surgery clinics struggling to assess new 'non-urgent' cases within 1-2 years, with these cases making up around half of all cases referred to shoulder surgery clinics. In many cases these patients have not tried effective non-surgical care, and while they wait 1-2 years to see a surgeon, they often remain untreated and develop symptoms which are complex and costly to manage.*

*A promising solution is a triage and management virtual service where physiotherapists screen 'non-urgent' referrals to shoulder surgery clinics and determine their suitability for non-surgical management (proposed new model of care). Similar models have been evaluated in some shoulder surgery clinics in Australia. For example, a hospital in Sydney using a similar model found that the waiting list was reduced by 34% and non-urgent musculoskeletal condition could be appropriately managed by a physiotherapist without needing to see a surgeon. Unfortunately, this type of model of care has not been evaluated in a proper clinical trial in Australia and no one has looked at whether a virtual service affects equitable access.*

*Our team is exploring people's perceptions of a new model of care investigating a V*IRtual physiotherapist-led Triage of referrals to shoUlder surgEons (VIRTUE) model of care *in public hospital surgical clinics for adults with rotator cuff-related shoulder pain referred to shoulder surgery clinics. This proposed new model of care – which is a term that describes how health services are organised and delivered – is based on a similar model for patients with low back pain referred for spine surgery which has been co-designed with several patients with low back pain and clinicians who manage people with low back pain.*

**Appendix 2. Interview and workshop guides**

*We believe that the proposed model of care could help people referred to a shoulder surgery clinic to receive effective care sooner, reducing the need for surgery in some cases.*

*Your input on this new model of care will help us further co-design and refine it before we evaluate it in a clinical trial.*

1. *Thank participants for their time. Get participant's consent and confirm that it is OK to audio record the interview. If yes, indicate when pressing record. Advise that:*
   a. *They can stop at any time, and continue later or not, whatever suits best.*
   b. *All responses will be anonymous and confidential.*
2. *Any questions before getting started?*

**Informant interview questions**
Listed below are a series of questions that may be asked to participants. They may be modified in response to early data collection to better explore emergent concepts raised by interviewees.

**a) <u>PATIENT INTERVIEW</u>**

**Section 1: Experiences with rotator cuff-related shoulder pain (CFIR 2 & 4)**

1. **Can you briefly describe your experience with rotator cuff-related shoulder pain ?**
   - When did you first experience it?
   - How does it affect your daily life and activities?
   - If you could go back to when your pain first started, is there anything you wish you'd known or done differently?
   - What would you want a healthcare provider or policymaker to truly understand about living with rotator cuff-related shoulder pain?
2. **What treatments or interventions have you tried for your rotator cuff-related shoulder pain?**
   - Have you seen a physiotherapist or explored other non-surgical options before considering surgery?
   - How effective did you find these treatments?
3. **What challenges have you faced in managing your rotator cuff-related shoulder pain?**
   - For example, access to care, affordability, understanding of treatment options, etc.
   - What would have made it easier to manage your rotator cuff-related shoulder pain or access care?
   - Have you ever felt dismissed or not believed by health professionals or others when talking about your pain?

**Appendix 2. Interview and workshop guides**

**Section 2: Experiences with the Referral to Shoulder Surgery Clinic [if applicable] CFIR 2 & 3/4)**

4. **Can you walk me through the process of being referred to a shoulder surgery clinic?**
    - Who referred you? (e.g., GP, specialist)
    - What was your understanding of why you were referred?
    - Were you involved in the decision to be referred?
5. **How did you feel about the referral process?**
    - Were there any parts of the process that were confusing, stressful, or unclear?
    - Was it what you expected?
6. **What information did you receive about the referral and what to expect from the shoulder surgery clinic?**
    - Did you feel adequately informed about what would happen next?

---

**Section 3: Perceptions and Acceptability of the VIRtual physiotherapist-led Triage of referrals to shoUlder surgEons (VIRTUE) model of care**

*[Explain the model of care] Our proposed new model of care involves a senior physiotherapist assessing new referrals to shoulder surgery clinics. The physiotherapist will talk with each patient via Zoom or similar to assess their medical history and needs, and then discuss the patient's care with the Surgical Team. Together, they will decide whether that patient would benefit most from non-surgical physiotherapist-led care or whether they need to see a shoulder surgeon. Patients assessed as needing to see a surgeon will be put on the Shoulder Surgery Clinic waiting list for the next available appointment. Patients who are suitable for non-surgical care will receive physiotherapist-led care as needed (this may be an in-person or via Zoom based on the patient's preference and to improve access in rural/remote areas). The patient will receive physiotherapist-led care typically provided in NSW Public Hospitals (e.g., advice to support self-management, exercise therapy). Following this period of physiotherapist-led care, the physiotherapist will consult with the Surgical Team again to determine if the patient should see a shoulder surgeon. Patients who may benefit from seeing a shoulder surgeon will be placed on the Shoulder Surgery Clinic waiting list. Patients who do not need to see a shoulder surgeon will be discharged from the Shoulder Surgery Clinic with a program for ongoing self-management provided by the physiotherapist.*

7. **What do you think about the idea of having physiotherapy assessment and treatment via Zoom or similar before seeing a shoulder surgeon? (TFA1)**

    - Do you think this is a good way to triage and manage patients, or would you prefer to see a surgeon directly first or a physiotherapist in-person first?
    - What makes you feel that way?
    - What do you think about the ease of accessing a computer or smartphone with internet at home to participate in virtual consultations?

8. **How would you feel about the idea of receiving physiotherapist-led care while you are waiting to see a surgeon (if surgery is still a possibility)? (TFA1 & 6)**
    - Does this approach seem helpful or potentially frustrating?

  - Would knowing that you will have physiotherapist-led care in the meantime change how you feel about waiting for surgery?
  - Do you think receiving physiotherapist-led care during the waiting period could help improve your condition, mental health or possibility prevent you from having surgery?

9. **What are your thoughts on the use of Zoom or similar (virtual consultations) for physiotherapy treatment, especially if you are on a waiting list to see a surgeon? (TFA1, 3 & 6)**
   - Do you feel comfortable using Zoom or similar for your consultation?
   - Do you think virtual physiotherapy appointments are as effective as in-person physiotherapy appointments?
10. **Would you feel confident in the recommendations made by a physiotherapist regarding whether you should continue with physiotherapist-led care or see a surgeon, even if you haven't yet seen a surgeon? (TFA1 & 6)**
    - Why or why not?
    - What would help you feel more confident in their recommendations?
    - Do you trust that the physiotherapist can make the right decision about your treatment without the surgeon's input yet? Does this cause any fear or anxiety?
11. **Our idea for how this proposed model of care might work is that the physiotherapist who does the triage evaluation will discuss each case with the surgeon before any final decisions about whether the patient needs to see a shoulder surgeon. What do you think about this approach? (TFA6)**
    - Do you think that this collaborative approach between physiotherapists and surgeons would be a better way to make treatment decisions?
    - Does knowing that both the physiotherapist and surgeon are involved in decision-making make you feel more confident in the process?
12. **The idea and initial design of our proposed model of care was inspired from another model which was developed in consultation with patients who have low back pain and clinicians who treat them. With that in mind, would this affect your opinion about its value and likely effectiveness? (TFA6)**
13. **What improvements, if any, would you suggest for this proposed new model of care to make it more acceptable, effective and efficient for people with rotator cuff-related shoulder pain? (TFA1)**
    - Are there any aspects of the process that you think could be streamlined, made more accessible, or improved in terms of support?

---

**Section 5: Closing Questions**

14. **Is there anything else you would like to share about your experience with rotator cuff-related shoulder pain, the referral process, or our ideas for a the VIRtual physiotherapist-led Triage of referrals to shoUlder surgEons (VIRTUE) model of care?**
    - Any additional feedback, suggestions, or concerns?

---

**Appendix 2. Interview and workshop guides**

**b) <u>CLINICIAN INTERVIEW</u>**

**Section 1: Experiences with rotator cuff-related shoulder pain management (CFIR 3 & 4)**

1. **Can you describe your experience in managing patients with rotator cuff-related shoulder pain?**
   - What are some of the challenges you commonly face in treating this condition? For example, issues with access to care, affordability, patient adherence, etc.
2. **What treatments or interventions do you commonly recommend to patients with rotator cuff-related shoulder pain?**
   - How effective have you found these treatments to be in your practice?
3. **What role (if any) does collaboration with other clinicians (e.g., physiotherapists, surgeons) play in your management of people with rotator cuff-related shoulder pain?**

---

**Section 2: Experiences with Referrals to Shoulder Surgery Clinics [if applicable] CFIR 3 & 4**

4. **Can you describe your experience with referring patients to shoulder surgery clinics?**
   - What factors influence your decision to refer a patient for surgery?
5. **How do you feel about the referral pathway to shoulder surgery clinics for rotator cuff-related shoulder pain patients?**
   - Are there any aspects of the process that you find confusing, frustrating, or unclear?
6. **What information do you typically provide to patients when referring them to a shoulder surgery clinic?**
   - Do you think patients are generally well-informed about the next steps and the purpose of the referral?

---

**Section 3: Perceptions and Acceptability of the VIRtual physiotherapist-led Triage of referrals to shoUlder surgEons (VIRTUE) model of care**

*[Explain the model of care] Our proposed new model of care involves a senior physiotherapist assessing new referrals to shoulder surgery clinics. The physiotherapist will talk with each patient via Zoom or similar to assess their medical history and needs, and then discuss the patient's care with the Surgical Team. Together, they will decide whether that patient would benefit most from non-surgical physiotherapist-led care or whether they need to see a shoulder surgeon. Patients assessed as needing to see a surgeon will be put on the Shoulder Surgery Clinic waiting list for the next available appointment. Patients who are suitable for non-surgical care will receive physiotherapist-led care as needed (this may be an in-person or via Zoom based on the patient's preference and to improve access in rural/remote areas). The patient will receive physiotherapist-led care typically provided in*

**Appendix 2. Interview and workshop guides**

*NSW Public Hospitals (e.g., advice to support self-management, exercise therapy). Following this period of physiotherapist-led care, the physiotherapist will consult with the Surgical Team again to determine if the patient should see a shoulder surgeon. Patients who may benefit from seeing a shoulder surgeon will be placed on the Shoulder Surgery Clinic waiting list. Patients who do not need to see a shoulder surgeon will be discharged from the Shoulder Surgery Clinic with a program for ongoing self-management provided by the physiotherapist.*

7. **What do you think about the idea of having a physiotherapist leading the triage process for rotator cuff-related shoulder pain patients via telehealth before they see a surgeon? TFA1**
    - Do you think this is a good way to manage referrals (virtual triage/consultation), or would you prefer that patients see a surgeon directly first or a physiotherapist in-person first?
8. **How do you think physiotherapists' involvement in triaging and managing patients before they see a shoulder surgeon could impact the healthcare system? TFA5 TFA6**
    - What outcomes would you want to see measured to determine if this model is successful?
    - What do you think are the main benefits or risks of having physiotherapists leading the triage and management process for rotator cuff-related shoulder pain patients? Could it reduce waiting times?

9. **In your opinion, how effective is telehealth for physiotherapy consultations compared to in-person physiotherapy consultations? TFA1 CFIR3**
    - Are there particular challenges or benefits you see with physiotherapist-led care via telehealth for patients with rotator cuff-related shoulder pain?
    - Have you used telehealth in your own practice for managing patients with musculoskeletal conditions?
10. **What improvements, if any, would you suggest to enhance the implementation/adoption of the proposed new model of care using a VIRtual physiotherapist-led Triage of referrals to shoUlder surgEons (VIRTUE) model of care? CFIR1**
    - How do you see the role of physiotherapists evolving within the multidisciplinary teams under this model?
    - In your opinion, what could improve the effectiveness or efficiency of the service?

11. **In your opinion, how could this service be integrated into current clinical workflows and improve patient care? CFIR 3 & 5**
    - What do you think would be the main challenges/barriers in integrating this new model of care into existing clinical workflows?
    - What would you propose to address these challenges/barriers?
    - What specific skills or training would be most important for physiotherapists to effectively evaluate and triage patients for surgical or non-surgical care?

**Appendix 2. Interview and workshop guides**

**Section 4: Closing Questions**

12. **Is there anything else you would like to share about your experiences in managing patients with rotator cuff-related shoulder pain, working with them, or your thoughts on the new VIRtual physiotherapist-led Triage of referrals to shoUlder surgEons (VIRTUE) model of care?**
    - Any additional feedback, suggestions, or concerns?

#### c. KEY STAKEHOLDER INTERVIEW

**Section 1: Organisational Experience with rotator cuff-related shoulder pain management (CFIR 3 & 4)**

1. **Can you describe your organisation's role in the management of patients with rotator cuff-related shoulder pain?**
    - What are the main services, policies, or initiatives your organisation offers that impact people with rotator cuff-related shoulder pain?
    - Does your organisation collaborate with other stakeholders (e.g., healthcare providers, government bodies, insurance companies) in the management of people with rotator cuff-related shoulder pain?
2. **What challenges does your organisation face on the management of people with rotator cuff-related shoulder pain at the healthcare system or population level?**
    - For example, challenges/barriers related to access to care, affordability, public awareness, or patient adherence.
3. **What kind of information does your organisation rely on to make decisions about the management of people with rotator cuff-related shoulder pain?**
    - How do you use this information to guide institution’s policies and/or intervention strategies?

---

**Section 2: Perspectives on the Referral Process to Shoulder Surgery Clinics (CFIR 3 & 4)**

4. **How do you feel about the current referral process to shoulder surgery clinics for rotator cuff-related shoulder pain patients?**
    - Are there any aspects of the referral process that you think need improvement? (e.g., timing, access, clarity for patients, coordination between providers)
    - Does your organisation have any policies, recommendations, or guidelines about when and how people should be referred to a shoulder surgery clinic?
5. **How does your organisation view the role of physiotherapy and non-surgical treatments in managing people with rotator cuff-related shoulder pain prior to referral to surgery?**
    - Do you have any policies, recommendations, or initiatives in place to promote physiotherapy as a first-line assessment and management of patients with rotator cuff-related shoulder pain?

**Appendix 2. Interview and workshop guides**

---

**Section 3: Perceptions and Acceptability of the VIRtual physiotherapist-led Triage of referrals to shoUlder surgEons (VIRTUE) model of care**

*[Explain the model of care] Our proposed new model of care involves a senior physiotherapist assessing new referrals to shoulder surgery clinics. The physiotherapist will talk with each patient via Zoom or similar to assess their medical history and needs, and then discuss the patient's care with the Surgical Team. Together, they will decide whether that patient would benefit most from non-surgical physiotherapist-led care or whether they need to see a shoulder surgeon. Patients assessed as needing to see a surgeon will be put on the Shoulder Surgery Clinic waiting list for the next available appointment. Patients who are suitable for non-surgical care will receive physiotherapist-led care as needed (this may be an in-person or via Zoom based on the patient's preference and to improve access in rural/remote areas). The patient will receive physiotherapist-led care typically provided in NSW Public Hospitals (e.g., advice to support self-management, exercise therapy). Following this period of physiotherapist-led care, the physiotherapist will consult with the Surgical Team again to determine if the patient should see a shoulder surgeon. Patients who may benefit from seeing a shoulder surgeon will be placed on the Shoulder Surgery Clinic waiting list. Patients who do not need to see a shoulder surgeon will be discharged from the Shoulder Surgery Clinic with a program for ongoing self-management provided by the physiotherapist.*

6. **What do you think about the idea of having a physiotherapist leading the evaluation using a triage process for patients with rotator cuff-related shoulder pain via telehealth before they see a shoulder surgeon? (TFA1)**
    - Do you have any concerns about this proposed model of care?
    - Would you support or promote this model of care within your organisation?
    - Do you believe telehealth led by a physiotherapist can be as effective as in-person physiotherapist-led care for managing people with rotator cuff-related shoulder pain? Why or why not?
7. **What impact do you think the introduction of a VIRtual physiotherapist-led Triage of referrals to shoUlder surgEons (VIRTUE) model of care could have on the broader healthcare system? (TFA5)**
    - Do you see it as a potential solution for reducing waiting times, improving access to care, or enhancing the overall patient experience? Why or why not?
    - How might your organisation's policies align with or support this new model of care?
8. **How do you think the integration of a physiotherapist-led evaluation using a triage process might affect the coordination of care and communication between healthcare providers? (TFA1 & 2)**
    - Do you think this collaborative model between physiotherapists and surgeons could improve the decision-making process for patient management?
9. **Are there any potential barriers or challenges that you see for health services in implementing this VIRtual physiotherapist-led Triage of referrals to shoUlder surgEons (VIRTUE) model of care? (TFA1, 2 & 5)**
    - For example, technological barriers, policy changes, financial constraints, or staff training.

10. **What improvements, if any, would you suggest for this new model of care to make it more effective or easier to implement at the organisational level? (TFA1, 2, & 4)**
    - Are there any adjustments that would make it more feasible or acceptable to stakeholders, including patients, healthcare providers, or insurance companies (for example)?

---

**Section 4: Closing Questions**

11. **Is there anything else you would like to share about your organisation's involvement in managing patients with rotator cuff-related shoulder pain or your thoughts on the new VIRtual physiotherapist-led Triage of referrals to shoUlder surgEons (VIRTUE) model of care?**
    - Any additional feedback, concerns, or suggestions for improving this model or the management of rotator cuff-related shoulder pain overall?

***END OF INTRVIEW GUIDES***

**Appendix 2. Interview and workshop guides**

**Stage 2: Co-design workshop guides:**

***Note: The topics below will serve as an outline to guide the workshops.***

**Introduction**

Introduce yourself to the workshop group.

Thank you for agreeing to participate in today's workshop. This workshop is voluntary and at any point of time you are free to withdraw. The workshop can take a maximum of 1-2 hours of your time today.

The purpose of this workshop is to codesign the **Co-Design of the** VIRtual physiotherapist-led Triage of referrals to shoUlder surgEons (VIRTUE) model of care before it is implemented in a trial.

3. Thank participants for their time. Check that it is OK to audio record the workshop. If yes, indicate when pressing record. Advise that:
   d. They can stop at any time, and continue later or not, whatever suits best.
   e. All responses will be anonymous and confidential.
4. Any questions before getting started?

Note for Facilitator: Follow-up prompts will be used to explore responses. For example, "What makes you say that?" or "Could you elaborate further?" This will allow an in-depth understanding of topics and responses. As indicated following each question, more specific prompts will be used to ensure complete coverage of topics raised, ensuring that workshops are in-depth and provide relevant data.

**Informant workshop questions**

Listed below are a series of discussion prompts and activities that may be asked to participants.

**Appendix 2. Interview and workshop guides**

#### <u>WORKSHOP 1: Patient Representatives Workshop</u>

Objective: Gather patient perspectives on the qualitative study findings and proposed model of care.

Structure:

Introduction (20 mins): Present study results and explain the purpose of the workshop.

- Welcome participants and introduce facilitators.
- Explain the purpose of the workshop and how their input will shape the model of care.
- Present key findings from the qualitative study.
- Provide a brief overview of the proposed model of care.

Discussion Group (40 mins):

- Reflections on Patient and Clinician Feedback from the qualitative study:
  - What are your thoughts on the concerns raised by the patient interviews?
  - What are your thoughts on the concerns raised by the clinician interviews?
- Model of Care Review:
  - Do you see your experiences reflected in the proposed model?
  - Are there any missing elements that should be included?
  - Are there any aspects that seem unnecessary or impractical?

Group Feedback Session (40 mins): Consolidate key themes from the discussion.

Wrap-Up & Next Steps (20 mins): Summarise insights and explain the next steps.

- Explain how their feedback will be incorporated into the next phase.
- Invite any final comments or questions.

**Appendix 2. Interview and workshop guides**

**WORKSHOP 2: Clinician and Key Stakeholder Representatives Workshop**

Objective: Gather clinician and key stakeholder perspectives on the qualitative study findings and proposed model of care.

Structure:

Introduction (20 mins): Recap patient feedback and qualitative study outcomes.

- Welcome clinicians and key stakeholders and introduce facilitators.
- Present key findings from the qualitative study.
- Provide an overview of the proposed model and its key components.

Discussion Group (40 mins):

- Reflections on Patient and Clinician Feedback from the qualitative study:
  - What are your thoughts on the concerns raised by the patient interviews?
  - What are your thoughts on the concerns raised by the clinician interviews?
  - How do these findings compare with your clinical experience?
- Model of Care Review:
  - What aspects of the model align with best clinical practices?
  - Are there any components that might be difficult to implement?
  - What additional resources or training would be needed?
  - What would make this model more practical in clinical settings?

Group Feedback Session (40 mins): Consolidate key themes from the discussion.

Wrap-Up & Next Steps (20 mins):

- Explain how their feedback will be incorporated into the next phase.
- Invite any final comments or questions.

**Appendix 2. Interview and workshop guides**

**WORKSHOP 3: Final Consensus Workshop (Patients, Clinicians and Key Stakeholders)**

Objective: Bring together patient, clinician and key stakeholder feedback to finalise the model of care.

Structure:

Recap (30 mins):

- Summarise key takeaways from Workshops 1 & 2.
- Present an updated version of the model based on prior feedback.

Collaborative Discussion (60 mins):

- Final Model Review:
  - Does the revised model address the key concerns raised in previous workshops?
  - Are there any remaining gaps?
  - What aspects of the model are most important to keep?
  - How can we ensure the model is both patient-centred and clinically feasible?

Consensus-Building Exercise (60 mins):

- Prioritise final changes using dot voting or a structured decision-making tool.
- Discuss the results and finalise key components.

Conclusion (30 mins):

- Confirm final agreements on the model of care.
- Thank participants and outline next steps – finalising the model report and commencement of VIRTUE trial.
